# Integrative analyses identify potential key genes and calcium-signaling pathway in familial AVNRT using whole-exome sequencing

**DOI:** 10.1101/2022.03.01.22271698

**Authors:** Jichang Huang, Rong Luo, Chenqing Zheng, Xin Cao, Yuncai Zhu, Tao He, Mingjiang Liu, Zhenglin Yang, Xiushan Wu, Xiaoping Li

**Affiliations:** Institute of Geriatric Cardiovascular Disease, Chengdu Medical College, Chengdu, People’s Republic of China; State Key Laboratory of Biocontrol, School of Life Sciences, Sun Yat-sen University, Guangzhou, China; School of Acupuncture-Moxibustion and Tuina, Chengdu University of Traditional Chinese Medicine, Chengdu, China; Department of Cardiology, Hospital of the University of Electronic Science and Technology of China and Sichuan Provincial People’s Hospital, Chengdu, Sichuan, China; The Center for Heart Development, Hunan Normal University, Changsha, Hunan, China

**Keywords:** Familial AVNRT, Arrhythmia, Whole-exome sequencing, Pathogenic genes, Calcium-signaling pathway

## Abstract

**Background:** Atrioventricular nodal reentrant tachycardia (AVNRT) is a common arrhythmia. Growing evidence suggests that family aggregation and genetic factors are involved in AVNRT. However, in families with a history of AVNRT, disease-causing genes have not been reported.

**Objective:** To investigate the genetic contribution of familial AVNRT using a WES approach.

**Methods:** Blood samples were collected from 20 patients from nine families with a history of AVNRT and 100 control participants, and we systematically analyzed mutation profiles using whole-exome sequencing. Gene-based burden analysis, integration of previous sporadic AVNRT data, pedigree-based co-segregation, protein-protein interaction network analysis, single-cell RNA sequencing, and confirmation of animal phenotype were performed.

**Results:** Among 95 reference genes, seven pathogenic genes have been identified both in sporadic and familial AVNRT, including *CASQ2, AGXT, ANK2, SYNE2, ZFHX3, GJD3*, and *SCN4A*. Among the 37 reference genes from sporadic AVNRT, five pathogenic genes were identified in patients with both familial and sporadic AVNRT: *LAMC1, RYR2, COL4A3, NOS1*, and *ATP2C2*. Considering the unique internal pathogenic gene within pedigrees, three genes, *TRDN, CASQ2*, and *WNK1*, were likely to be the pathogenic genes in familial AVNRT. Notably, the core calcium-signaling pathway may be closely associated with the occurrence of AVNRT, including *CASQ2, RYR2, TRDN, NOS1, ANK2*, and *ATP2C2*.

**Conclusion:** These results revealed the underlying mechanism for familial AVNRT.

## Introduction

Atrioventricular nodal reentrant tachycardia (AVNRT) is a relatively common arrhythmia, accounting for approximately 45%-65% of paroxysmal supraventricular tachycardia (PSVT).^1^ The heart rate of a normal adult is typically 60 to 100 beats per minute, whereas the heart rate of patients with AVNRT exceeds 150 beats per minute.^2-4^ This continuous re-excitement of the myocardium can induce arrhythmias, syncope, and even sudden death.

Slow and fast atrioventricular nodal pathways are currently recognized as the pathobiological mechanism for AVNRT, wherein the calcium-signaling pathway may be a crucial regulator.^5^ Calmodulin-dependent protein kinase II (CaMKII) can directly phosphorylate L-type voltage-gated calcium channels (Cav1.2) to increase Ca^2+^ influx in cardiomyocytes,^6^ inducing early depolarization and causing arrhythmia.^7^ In addition, CaMKII can phosphorylate the ryanodine receptor 2 (*RYR2*) on the sarcoplasmic reticulum (SR) to release a large amount of Ca^2+^ into the cytoplasm from SR.^8^ Excessive Ca^2+^ activates the Na^+^/Ca^2+^ exchanger (NCX), resulting in spontaneous myocyte depolarization and abnormal rhythm.^8^ Furthermore, the inhibition of NO synthase 1 (NOS1) in SR decreased *RYR2* activity because of reducing Ca^2+^ sparks and shortened action potential causing arrhythmia susceptibility.^9^ Although radiofrequency ablation for the treatment of AVNRT has shown good results, its precise reentry path and its molecular mechanism remain to be explained.

AVNRT was considered a sporadic disease in the past, with a prevalence of 22.5 cases per 10,000 persons.^10^ Nevertheless, several studies have reported that AVNRT occurred in twins and the same family member,^10-13^ indicating the phenomenon of family clustering of AVNRT. To date, few studies of AVNRT pedigrees have been available,^10, 12, 14, 15^ as this is relatively a rare phenomenon. Familial AVNRT pedigree was reported for the first time in 2004.^12^ Subsequently, the European clinical study reported 24 AVNRT pedigrees in 2017.^10^ Recently, we described the clinical reports of eight families with a history of AVNRT in China in 2021.^15^

The familial AVNRT phenomenon indicates that genetic factors play a crucial role in AVNRT pathogenesis; however, investigations at a molecular level are currently lacking. No report is available on the pathogenic genes of AVNRT. In addition, only two studies have explored the screening of pathogenic genes of AVNRT.^5, 16^ In 2018, Andreasen et al. first sequenced 67 known pathogenic genes associated with arrhythmia in 298 patients with AVNRT and reported mutations in genes encoding various Na^+^ and Ca2^+^ channels,^16^ suggesting that AVNRT is associated with various ion channels. Recently, we found that AVNRT is closely associated with the neuronal system or ion channels, and 10 potential candidate pathogenic genes were screened out in 82 patients with sporadic AVNRT using whole-exome sequencing (WES).^5^ Although variants of genes were identified in patients with sporadic AVNRT, it is difficult to identify the disease phenotype and genotype. Fortunately, the emergence of pedigree-based studies addressed this issue.^17^ The pedigree-based study had several advantages for a rare variant: reduced genetic heterogeneity, enriched rare alleles, and co-segregated with the disease phenotype and genotype.^18^ Therefore, we hypothesized that the application of a more integrated approach might help elucidate the genetic etiology of AVNRT disease.

To the best of our knowledge, this is the first study that primarily aimed to investigate the genetic contribution of familial AVNRT using a WES approach. In this study, we used WES to identify potential key genes on the basis of gene-based burden, pedigree-based co-segregation, protein-protein interaction (PPI) analyses, single-cell RNA sequencing, and confirmation of phenotype for AVNRT disease.

## Methods

### Collection of peripheral blood samples

Patients with AVNRT were enrolled in the Sichuan Provincial People’s Hospital in China from 2013 to 2020. Familial AVNRT defined that two or more AVNRT patients in a family, or 1 or more clinically diagnosed PSVT patients in a family of AVNRT proband patient. In addition, 100 unrelated ethnically matched healthy participants were recruited from the Sichuan Provincial People’s Hospital. All patients were diagnosed with AVNRT using radiofrequency catheter ablation. Healthy participants did not have a history of cardiovascular diseases, arrhythmia, systemic immune diseases, cancers, or any other diseases known to cause arrhythmias. Whole blood samples from 20 patients with AVNRT and 100 normal control participants were collected in heparinized vacutainer tubes. Patients had signed an informed consent form before enrollment. This study was approved by the ethics committee of the Sichuan Academy of Medical Sciences and the Sichuan Provincial People’s Hospital.

### Intracardiac electrophysiological study

Intracardiac electrophysiology recordings included atrial stimulation (sudden or additional stimulation pacing) and ventricular stimulation in patients. AVNRT diagnosis is established on the basis of published standards and applicable pacing operations. The physiology of atrioventricular node is defined as the atrial-His (AH) interval increase of ≥50 ms after a decreasing interval of 10 ms during the additional stimulation of the single atrium or the AH interval increase of ≥50 ms after the pacing cycle length is shortened by 10 ms. If continuous AVNRT is not induced (lasting more than 30 s), the same pacing operation was repeated with isoproterenol administration and withdrawal as described above.

### Whole-exome sequencing, variant selection, and annotation

In brief, we purified DNA from the peripheral blood using the QIAamp DNA Blood Mini Kit (Qiagen, Hilden, Germany). Whole-exome enrichment was performed using the SureSelect Human All Exon kit V6 (Agilent Technologies, Santa Clara, CA, USA). The genomic DNA library was sequenced using the HiSeq X and NovaSeq systems (Illumina, San Diego, CA, USA). The sequenced DNA fragments were aligned with Human Reference Genome (National Center for Biotechnology Information Build 37) on the basis of the Burrows–Wheeler transform. The removal of duplication, realignment, and recalibration were performed with Picard tools (http://picard.sourceforge.net/) and GATK (http://www.broadinstitute.org/gsa/wiki/index.php/Home_Page). The single-nucleotide polymorphisms and insertion-deletion polymorphisms (indels) were performed using GATK3.7 software. The high-confidence variants were annotated with snpEff (Version 4.2; http://snpeff.sourceforge.net/). In addition, the annotations of all variants were further performed using 1000 Genomes Project data (2014 Oct release, http://www.1000genomes.org), the Exome Aggregation Consortium (http://exac.broadinstitute.org), EVS (http://evs.gs.washington.edu/EVS), the ClinVar (http://www.ncbi.nlm.nih.gov/clinvar) database, and Online Mendelian Inheritance in Man (http://www.omim.org).

### Rare variants of the pathogenic reference genes

In total, 95 reference genes were selected for the analysis of rare variants in patients with AVNRT and control participants. These genes were considered reference genes according to our previous study.^5^ To increase reliability and generalizability of reference genes, data integration was used to select the genes following our previous sporadic AVNRT study. Therefore, the reference genes were identified in patients with both the sporadic and familial AVNRT and were assessed for segregation within families.

Biological process (BP) of Gene Ontology (GO) was performed by database for annotation, visualization and integrated discovery (DAVID) bioinformatics resources according to previous study.^19^ Protein–protein interactions (PPI) network of candidate genes were obtained from the STRING database (https://cn.string-db.org). The images of single-cell sequencing data from healthy human cardiac tissue were obtained from the Human Protein Atlas (see http://www.proteinatlas.org). The mouse phenotypes associated with pathogenic reference genes were extracted from the Mouse Genome Informatics (MGI) database (http://www.informatics.jax.org/).

### Rare variants of common pathogenic genes in sporadic and familial AVNRT

To identify the common pathogenic mechanisms in all AVNRT cases, the 37 most likely pathogenic genes from our previously sporadic AVNRT study^5^ were considered the intersection of both sporadic and familial AVNRT, and then pathogenic genes were identified in patients with both familial and sporadic AVNRT, and were assessed for segregation within families.

BP analysis was performed by DAVID bioinformatics resources.^19^ The PPI network of candidate genes was obtained from the STRING database (https://cn.string-db.org). The images of single-cell sequencing data from healthy human cardiac tissue were obtained from the Human Protein Atlas (http://www.proteinatlas.org). The mouse phenotypes associated with pathogenic reference genes were extracted from the Mouse Genome Informatics (MGI) database (http://www.informatics.jax.org/).

### Rare variants of potential pathogenic genes in familial AVNRT

To analyze the aggregate association of rare variants at the gene level, we performed gene-based burden analysis to obtain gene-level significant associations of familial AVNRT patients (n = 20) and control subjects (n = 100). Rare variants were defined as ‘deleterious variants’ according to 1000 Genomes Project data and ExAC with MAF< 0.001, MAF< 0.01, or MAF< 0.05. Fisher’s exact test was used to evaluate gene-based burden analysis. The gene level across the genome was used to identify risk genes across different allele frequency spectrums.

The significant genes were submitted to the KOBAS3.0 web server (http://kobas.cbi.pku.edu.cn/kobas3) to obtain the functional gene set Reactome Pathway enrichment. The PPI network of candidate genes was obtained from the STRING database (https://cn.string-db.org). The images of single-cell sequencing data from healthy human cardiac tissue were obtained from the Human Protein Atlas (http://www.proteinatlas.org). The mouse phenotypes associated with pathogenic reference genes were extracted from the Mouse Genome Informatics (MGI) database (http://www.informatics.jax.org/).

### Protein–protein interactions network of potential pathogenic genes

Protein–protein interactions (PPI) network of candidate genes were obtained from the STRING database (https://cn.string-db.org). The relationships among the screened genes were predicted by STRING database and visualized with Cytoscape v2.3 software.

## Results

### Clinical data of the patients

In this study, a total of 20 patients and 100 control participants were included to perform WES. These 20 patients were assessed in nine families, including a total of 93 members. Among 20 patients enrolled in this study, the male to female ratio was 1.86, the mean age at onset was approximately 47.5 years, the heart rate at onset was approximately 176.9 beats per minute, and all the patients were free from structural heart disease (Table 1). All the patients showed typical slow-fast AVNRT, and 60.0% of the patients were successfully treated by radiofrequency ablation during the operation.

**Table 1.** Demographic baseline of patients.

| Variables | Total patients (n = 20) |
| --- | --- |
| Sex, male (%) | 13 (65.0) |
| Age at onset, year | 47.5 ±14.3 |
| Heart rate at onset, bpm | 176.9 ±12.8 |
| Structural heart disease, yes (%) | 0 (0) |
| AVNRT Type, typical (100%) | 20 (100) |
| Radiofrequency ablation, yes (%) | 12 (60.0) |

bpm: beat per minute.

### Rare variants of the pathogenic reference genes

To increase reliability and generalizability of reference genes, data integration was used to confirm the genes following our previous sporadic AVNRT study.^5^ Therefore, patients with both sporadic and familial AVNRT were enrolled in this study. Among the 95 reference genes, seven pathogenic genes have been identified in patients with both familial and sporadic AVNRT: *CASQ2, AGXT, ANK2, SYNE2, ZFHX3, GJD3*, and *SCN4A* (Table S1). Rare variants of *AGXT, ANK2, SYNE2, GJD3*, and *SCN4A* co-segregated within one pedigree and those of *CASQ2* and *ZFHX3* within two and three pedigrees, respectively (Table S1).

The bubble plot of GO-BP analysis showed that the functions of these genes were mainly associated with cardiac conduction, muscle contraction, and the release of sequestered calcium ions (Table S1 and Figure 1A). Furthermore, PPI networks of these genes indicated that *CASQ2, ANK2*, and *SCN4A* constituted the network, and *ZFHX3* interacted with *SYNE2* (Figure 1B). In addition, the results of single-cell sequencing showed that the relative expression of *CASQ2, ANK2*, and *SYNE2* was higher in cardiomyocytes than others (Figure 1C and Supplementary Figure 1), whereas the expression of *SCN4A, ZFHX3, AGXT*, and *GJD3* was relatively lower or not expressed (Figure 1C and Supplementary Figure 1).

**Figure 1.**
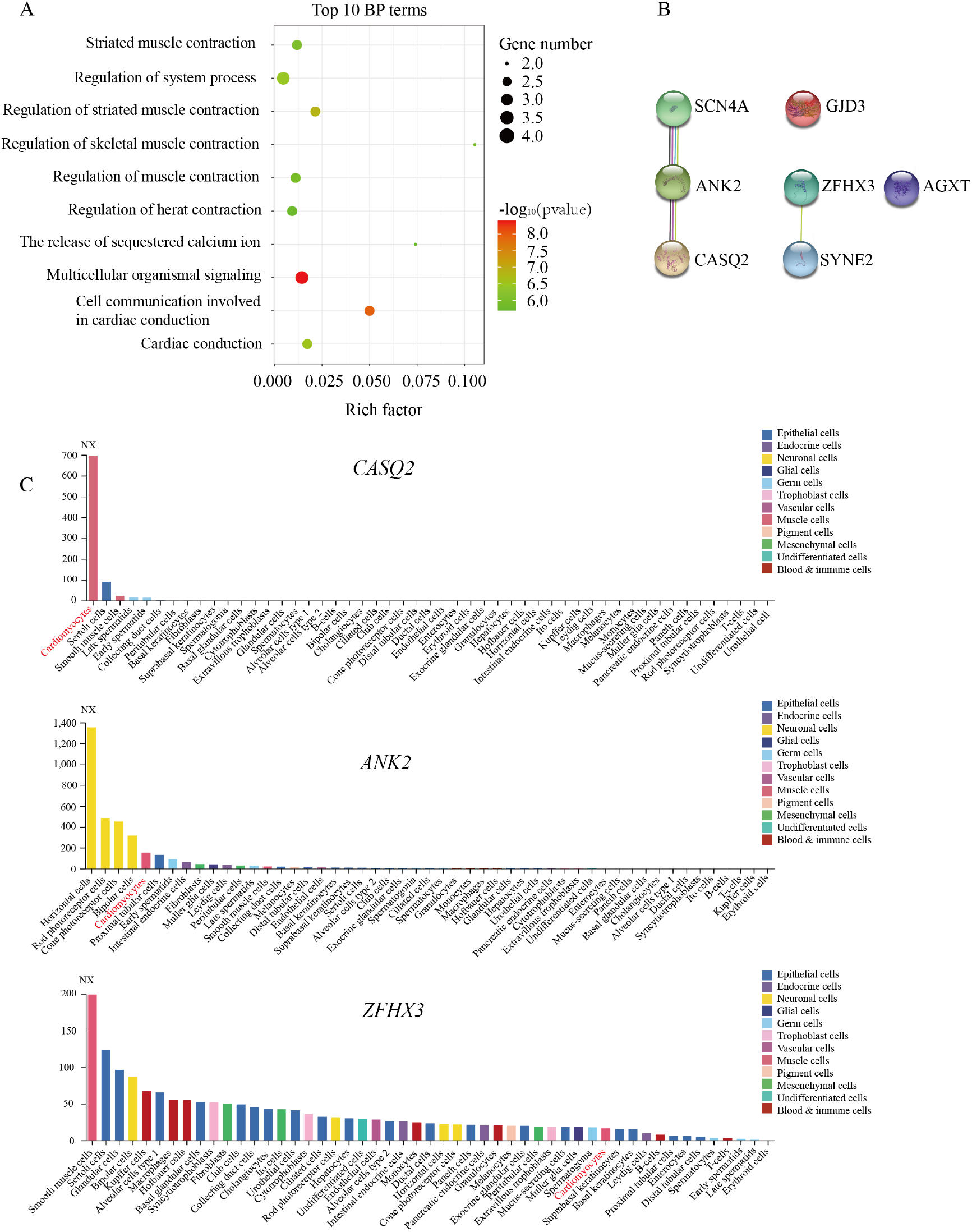
Identification of pathogenic reference genes in familial atrioventricular nodal reentrant tachycardia. (A) The top 10 biological process terms of seven pathogenic genes are depicted using enrichment analysis (P < 0.05). (B) The protein–protein interactions analysis of seven pathogenic genes. (C) The expression of *CASQ2, ANK2*, and *ZFHX3* was shown in different cell types by the single-cell sequencing data.

To further verify these gene functions, the Mouse Genome Informatics (MGI) database was used to confirm their phenotype. The disruption of *CASQ2*, essential for Ca^2+^ storage, led to ventricular tachycardia in both mice and humans (Table S1). Moreover, the abnormal function of ankyrin-2 (*ANK2*) may lead to sinoatrial node disease and ankyrin-B-related cardiac arrhythmia in humans (Table S1). In addition, its abnormality increased heart rate variability and caused the abnormal sinoatrial node conduction in the mouse (Table S1). *ZFHX3* was identified as a crucial risk factor for atrial fibrillation,^20^ *SYNE2* contributed to cardiac arrhythmia (Table S1), and *GJD3* caused abnormal atrioventricular node conduction (Table S1). In summary, in light of our analyses and previous study, we suggested that *CASQ2, ANK2*, and *ZFHX3* were the most likely pathogenic genes for AVNRT.

### Rare variants of common pathogenic genes in sporadic and familial AVNRT

To identify the common pathogenic mechanisms in all AVNRT cases, the 37 most likely pathogenic genes from our previous sporadic AVNRT study were considered the intersection of both sporadic and familial AVNRT. Among these genes, five pathogenic genes were identified in patients with both familial and sporadic AVNRT: *LAMC1, RYR2, COL4A3, NOS1*, and *ATP2C2* (Table S2). Furthermore, the rare variants of *LAMC1, COL4A3, NOS1*, and *ATP2C2* co-segregated within one pedigree apart from *RYR2* (Table S2).

The BP enrichment analysis suggested that the functions of these genes were mainly associated with heart contraction and the regulation of calcium ion (Figure 2A and Table S2). In addition, the PPI networks showed that *RYR2, NOS1*, and *ATP2C2* constituted the network, and *COL4A3* interacted with *LAMC1* (Figure 2B). Moreover, the results of single-cell sequencing data showed that the relative expression of *RYR2* and *LAMC1* was higher in cardiomyocytes than others (Figure 2C and Supplementary Figure 2), whereas the expression of *COL4A3, NOS1*, and *ATP2C2* was relatively lower or not expressed (Figure 2C and Supplementary Figure 2).

**Figure 2.**
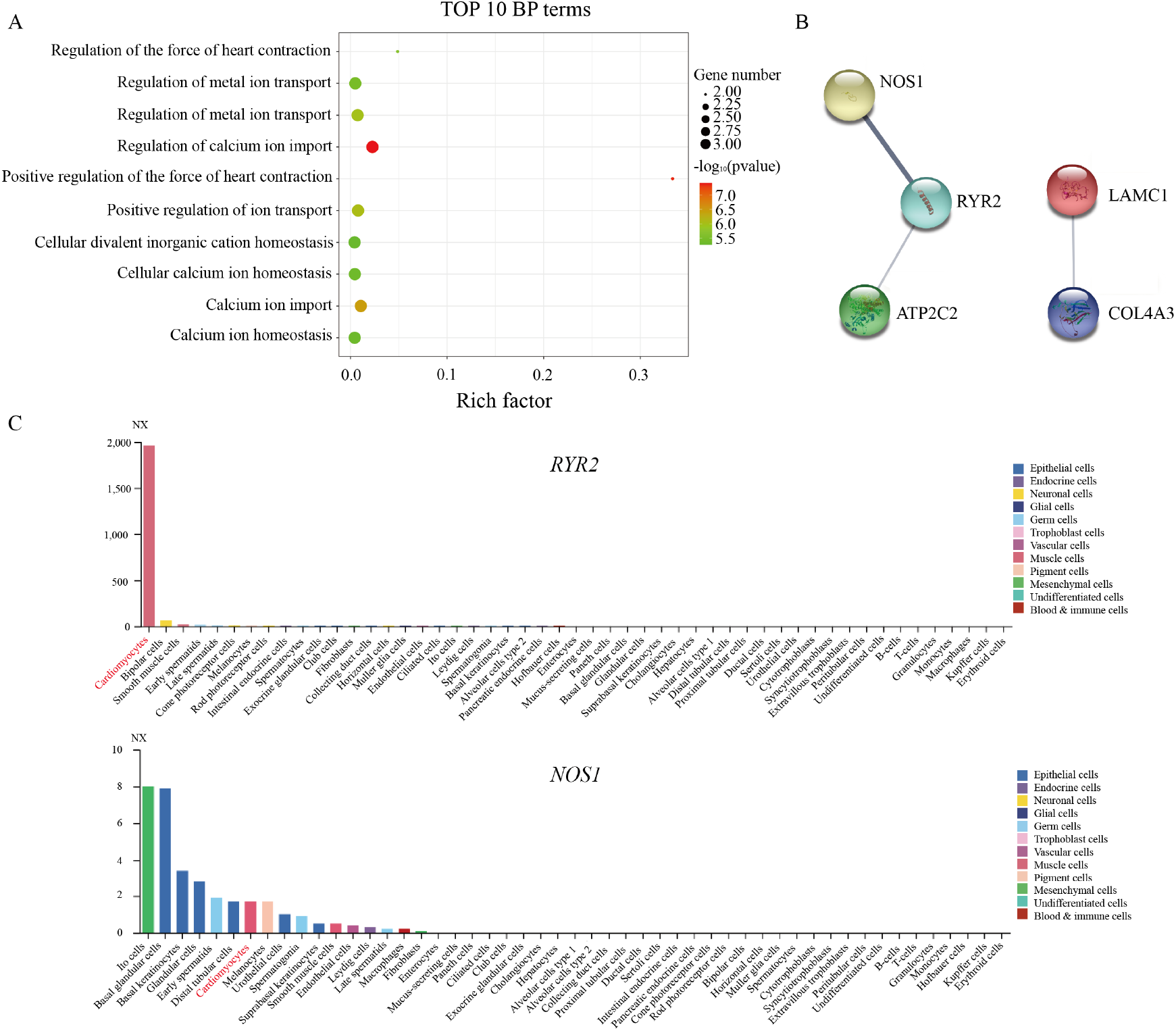
Identification of common pathogenic genes in sporadic and familial atrioventricular nodal reentrant tachycardia. (A) The top 10 biological process terms of five pathogenic genes are depicted using enrichment analysis (P < 0.05). (B) The protein–protein interactions analysis of five pathogenic genes. (C) The expression of *RYR2* and *NOS1* was shown in different cell types using the single-cell sequencing data.

The biological function and phenotype of these genes were further explored using the MGI database. Homozygous mutation in the *NOS1* gene led to abnormal cardiac muscle relaxation and increased heart rate in the mouse (Table S2). The disruption of *RYR2* was associated with arrhythmogenic right ventricular dysplasia and catecholaminergic polymorphic ventricular tachycardia in humans, whereas it is mainly associated with an increased heart rate and ventricular tachycardia in the mouse (Table S2). However, cardiac diseases were independent of the functions of *LAMC1, COL4A3*, and *ATP2C2* (Table S2). Considering their functions and previous study, *RYR2* and *NOS1* were likely to be causal genes for AVNRT.

### Rare variants of pathogenic genes in familial AVNRT

In search of the underlying pathogenic mechanisms within AVNRT pedigrees, we imposed more restrictive criteria: more than two mutations and one homozygous mutation in one gene segregated at least two pedigrees. A total of 299 genes with 452 rare variants were identified (Table S3).

As shown in Table S3, the three AVNRT-related traits among the pathways in the Reactome databases were as follows: (1) stimuli-sensing channels, (2) RYR tetramers transport Ca^2+^ from the SR lumen to the cytosol, and (3) ion channel transport. In addition, seven pathogenic genes were identified, including *TRDN, ANO6, SLC9C1, CASQ2, ATP6V0A4, SGK2*, and *WNK1*. Remarkably, *CASQ2* has been involved in AVNRT as reference genes.

MGI database and previous studies was further used to confirm the phenotype of these genes. The disruption of *TRDN* contributed to catecholaminergic polymorphic ventricular tachycardia in humans.^21^ Mice lacking *ANO6* developed shortened PQ intervals (Table S3). The aberration of *WNK1* led to hereditary sensory and autonomic neuropathy in humans.^22^ Considering their functions and previous study, *TRDN, CASQ2*, and *WNK1* were likely to be the common pathogenic genes in familial AVNRT.

### The calcium-signaling pathway of AVNRT

To explore the internal relationship of the candidate pathogenic genes in this study, PPI network analysis was further constructed. Among these 14 candidate pathogenic genes, three networks were established; these genes were *CASQ2, AGXT, ANK2, SYNE2, ZFHX3, GJD3, SCN4A, LAMC1, RYR2, COL4A3, NOS1, ATP2C2, TRDN*, and *WNK1* (Figure 4A). PPI networks indicated that the genes constituted network 1 (*CASQ2, ANK2, SCN4A, RYR2, NOS1, ATP2C2*, and *TRDN*), network 2 (*SYNE2* and *ZFHX3*), and network 3 (*LAMC1* and *COL4A3*).

The maximum network 1 was mainly associated with the calcium-signaling pathway using Kyoto Encyclopedia of Genes and Genomes enrichment analysis (Figure 3B). Among them, *RYR2* acted as a calcium channel that released calcium ions into the cytoplasm from the SR and thus regulated cardiac muscle contraction.^23^ The RYR forms a complex with *TRDN*, junction (*JTC*), and *CASQ* instead of acting independently.^24^ Moreover, the mutations of *RYR2* or *CASQ* lead to Ca^2+^ leak in ventricular tachycardia and thus contribute to Ca^2+^ waves in arrhythmogenic as a result of the increasing Ca^2+^ spark frequency and rising flux.^24, 25^ Particularly, *CASQ2* and *TRDN* have also been identified in this study. Furthermore, another *RYR2*-related gene is neuronal *NOS1*, which is located in the cardiac SR and enhances contraction through NO production.^26^ The present studies have shown that the inhibition of *NOS1* decreased *RYR2* activity because of reducing Ca^2+^ sparks and shortened action potential causing arrhythmia susceptibility.^9, 26^ In addition, *ANK2* from the SR promotes the flow of calcium ions into the plasma membrane through the inositol triphosphate receptor and NCX.^27^ *ATP2C2* encodes calcium-transporting ATPase, removing calcium from the cytosol into the Golgi body.^28^ Therefore, the calcium-signaling pathway may be closely associated with the occurrence of AVNRT.

**Figure 3.**
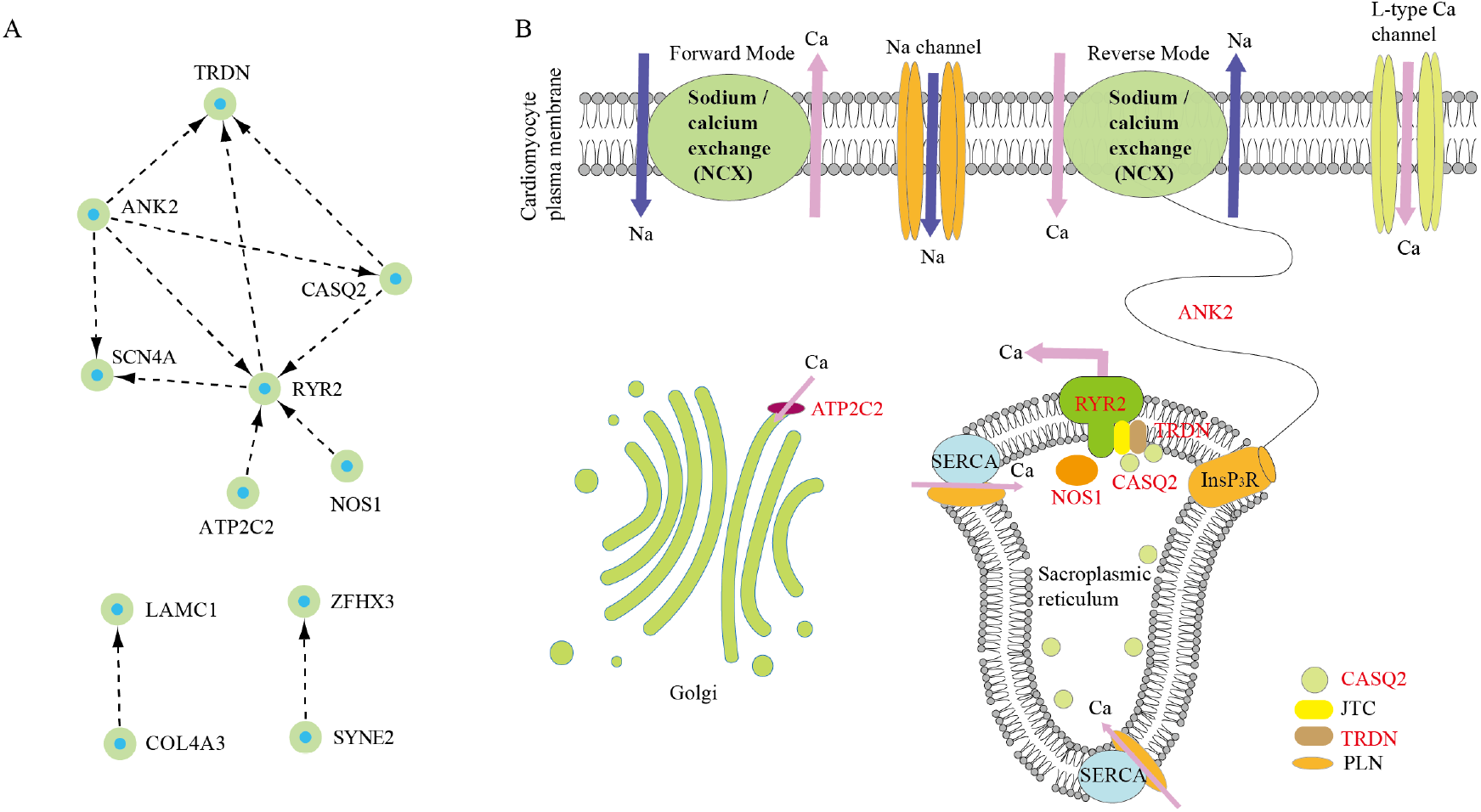
Identification of core-signaling pathway in atrioventricular nodal reentrant tachycardia (AVNRT). (A) The protein–protein interactions analysis of 14 pathogenic genes. (B) Schematic representation of core-signaling pathway in AVNRT.

## Discussion

Although significant inroads have been achieved in elucidating the pathogenesis of AVNRT,^1, 5^ the molecular mechanisms associated with this disease remain in its early stages. The sporadic studies contributed to the discovery of a large number of candidate pathogenic genes;^5^ however, it is difficult to effectively rule out unrelated genes. Unlike the sporadic studies, the pedigree-based linkage study directly observes the segregation of variants with disease phenotype.^17^ The integrated analysis of sporadic and familiar studies may provide novel strategies for exploring the more prevalent pathogenesis. Phenotypes associated with pathogenic genes were further confirmed using the MGI database. Thus, we took advantage of phenotype analysis and integrated sporadic and pedigree analyses to reveal the novel genetic associations with AVNRT. In this study, genes such as *CASQ2, ANK2, ZFHX3, RYR2, NOS1, TRDN*, and *WNK1* were likely pathogenic.

Recently, accumulating studies have revealed that genetic factors may contribute to the pathogenesis of AVNRT.^10-13^ However, little is known about the genetic role of AVNRT. In 298 patients with AVNRT, the disease was observed to be associated with Na^+^ and Ca2^+^ channels detected using next-generation sequencing.^16^ Recently, we, for the first time, found that AVNRT was closely associated with the neuronal system or ion channels, and 10 potential candidate pathogenic genes were screened out in 82 patients with sporadic AVNRT using WES.^5^ Among the pathogenic reference genes, multiple variants in ion channel genes *(CASQ2, ANK2*, and *SCN4A*) were further confirmed both in previous sporadic^5^ and these familial studies. The gene *CASQ2*, encoding the calcium-binding protein, played a crucial role in excitation-contraction coupling, regulated the heart rate, and was associated with ventricular tachycardia.^29-31^ Moreover, another calcium ion transport-related gene *ANK2* may lead to cardiac arrhythmia.^32^ *ZFHX3* was identified as a crucial risk factor for atrial fibrillation,^33^ and *SYNE2* contributed to atrial fibrillation.^34^ These results suggested calcium handling might have played a crucial role in the pathogenesis of AVNRT.

Slow and fast atrioventricular nodal pathways are currently recognized as the mainly pathobiological mechanism for AVNRT, wherein the calcium-signaling pathway may be a crucial regulator.^5^ CaMKII can directly phosphorylate L-type voltage-gated calcium channels (Cav1.2) to increase Ca^2+^ influx in cardiomyocytes, inducing early depolarization and causing an arrhythmia.^6^ Moreover, CaMKII can phosphorylate the *RYR2* on the SR to release a large amount of Ca^2+^ into the cytoplasm from the SR, and excessive Ca^2+^ activates the NCX, resulting in spontaneous myocyte depolarization and abnormal rhythm.^8^ Furthermore, the inhibition of *NOS1* in the SR decreased RYR2 activity because of reducing Ca^2+^ sparks and shortened action potential causing arrhythmia susceptibility.^9^

In both sporadic and familial AVNRT, there were six calcium channel-associated genes, including *RYR2, NOS1, TRDN, CASQ2, ANK2*, and *ATP2C2. RYR2* acted as a calcium channel that released calcium ions into the cytoplasm from the SR and thus regulated cardiac muscle contraction.^23^ The RYR forms a complex with *TRDN, JTC*, and *CASQ* instead of acting independently.^24^ Moreover, the mutations of *RYR2* or *CASQ* lead to Ca^2+^ leak in ventricular tachycardia, thus contributing to Ca^2+^ waves in arrhythmogenic as a result of the increasing Ca^2+^ spark frequency and rising flux.^24, 25^ Particularly, *CASQ2* and *TRDN* have also been identified in this study. Furthermore, another *RYR2*-related gene was neuronal *NOS1. NOS1* is located in the cardiac SR and enhances contraction through NO production.^26^ The present studies showed that the inhibition of *NOS1* decreased *RYR2* activity because of reducing Ca^2+^ sparks and shortened action potential causing arrhythmia susceptibility.^9, 26^ From these findings, *RYR2* as the core-signaling pathway may be closely associated with the occurrence of AVNRT. The functions of these calcium channel-associated genes are currently being explored in functional experiments.

To the best of our knowledge, this is the first study primarily aimed to investigate the genetic contribution of familial AVNRT using a WES approach. The calcium-signaling pathway should be considered seriously for AVNRT.

## List of abbreviations

AVNRT: Atrioventricular nodal reentrant tachycardia
SNPs: single nucleotide polymorphisms
indels: insertion-deletion polymorphisms
ExAC: Exome Aggregation Consortium
OMIM: Online Mendelian Inheritance in Man
CaMKII: Calmodulin-dependent protein kinase II
*RYR2*: ryanodine receptor 2
SR: sarcoplasmic reticulum
*NOS1*: NO synthase 1
WES: whole-exome sequencing
indels: insertion-deletion polymorphisms
STRING: Search Tool for the Retrieval of Interacting Genes database
GO: Gene Ontology
BP: Biological process
PPI: protein–protein interactions
MAF: minor allele frequency
DAVID: database for annotation, visualization and integrated discovery.

## Data availability statement

All data are incorporated into the article and its supplementary material.

## Conflict of Interest

None declared

## Funding

This work was supported by grants from Chinese National Natural Science Foundation (No. 81770379, 32171182, and 81670290), and the Foundation of Chengdu Medical College (CYZZD21-04, 2021LHPJ-02).

## Notes

### Competing Interest Statement

The authors have declared no competing interest.

### Author Declarations

The ethics committee of the Sichuan Academy of Medical Sciences and the Sichuan Provincial People's Hospital gave ethical approval for this work

## Reference

1. Akhtar M, Jazayeri MR, Sra J, Blanck Z, Deshpande S, Dhala A. Atrioventricular nodal reentry. Clinical, electrophysiological, and therapeutic considerations. Circulation Jul 1993;88:282–295.

2. Porter MJ, Morton JB, Denman R, Lin AC, Tierney S, Santucci PA, Cai JJ, Madsen N, Wilber DJ. Influence of age and gender on the mechanism of supraventricular tachycardia. Heart Rhythm Oct 2004;1:393–396.

3. Gonzalez-Torrecilla E, Almendral J, Arenal A, Atienza F, Atea LF, del Castillo S, Fernandez-Aviles F. Combined evaluation of bedside clinical variables and the electrocardiogram for the differential diagnosis of paroxysmal atrioventricular reciprocating tachycardias in patients without pre-excitation. J Am Coll Cardiol Jun 23 2009;53:2353–2358.

4. Liuba I, Jonsson A, Safstrom K, Walfridsson H. Gender-related differences in patients with atrioventricular nodal reentry tachycardia. Am J Cardiol Feb 1 2006;97:384–388.

5. Luo R, Zheng C, Yang H, Chen X, Jiang P, Wu X, Yang Z, Shen X, Li X. Identification of potential candidate genes and pathways in atrioventricular nodal reentry tachycardia by whole-exome sequencing. Clin Transl Med Jan 2020;10:238–257.

6. Cheng J, Xu L, Lai D, Guilbert A, Lim HJ, Keskanokwong T, Wang Y. CaMKII inhibition in heart failure, beneficial, harmful, or both. Am J Physiol Heart Circ Physiol Apr 1 2012;302:H1454–1465.

7. Weiss JN, Garfinkel A, Karagueuzian HS, Chen PS, Qu Z. Early afterdepolarizations and cardiac arrhythmias. Heart Rhythm Dec 2010;7:1891–1899.

8. Maier LS, Zhang T, Chen L, DeSantiago J, Brown JH, Bers DM. Transgenic CaMKIIdeltaC overexpression uniquely alters cardiac myocyte Ca2+ handling: reduced SR Ca2+ load and activated SR Ca2+ release. Circ Res May 2 2003;92:904–911.

9. Wang H, Viatchenko-Karpinski S, Sun J, Gyorke I, Benkusky NA, Kohr MJ, Valdivia HH, Murphy E, Gyorke S, Ziolo MT. Regulation of myocyte contraction via neuronal nitric oxide synthase: role of ryanodine receptor S-nitrosylation. J Physiol Aug 1 2010;588:2905–2917.

10. Michowitz Y, Anis-Heusler A, Reinstein E, Tovia-Brodie O, Glick A, Belhassen B. Familial Occurrence of Atrioventricular Nodal Reentrant Tachycardia. Circ Arrhythm Electrophysiol Feb 2017;10:e004680.

11. Lu CW, Wu MH, Chu SH. Paroxysmal supraventricular tachycardia in identical twins with the same left lateral accessory pathways and innocent dual atrioventricular pathways. Pacing Clin Electrophysiol Oct 2000;23:1564–1566.

12. Hayes JJ, Sharma PP, Smith PN, Vidaillet HJ. Familial atrioventricular nodal reentry tachycardia. Pacing Clin Electrophysiol Jan 2004;27:73–76.

13. Namgung J, Kwak JJ, Choe H, Kwon SU, Doh JH, Lee SY, Lee WR. Familial occurrence of atrioventricular nodal reentrant tachycardia in a mother and her son. Korean Circ J Oct 2012;42:718–721.

14. Stec S, Deutsch K, Zienciuk-Krajka A. The world’s largest family with familial atrio-ventricular nodal reentry tachycardia. Kardiol Pol 2015;73:1339.

15. Chen XP, Yan C, Luo R, Zhu Y, Qian M, Liu X, Liu M, Ikeda T, Li X. Clinical report of 8 families with atrioventricular nodal reentrant tachycardia from China. Kardiol Pol Feb 25 2021;79:185–187.

16. Andreasen L, Ahlberg G, Tang C, et al. Next-generation sequencing of AV nodal reentrant tachycardia patients identifies broad spectrum of variants in ion channel genes. Eur J Hum Genet May 2018;26:660–668.

17. Wolf JB, Lindell J, Backstrom N. Speciation genetics: current status and evolving approaches. Philos Trans R Soc Lond B Biol Sci Jun 12 2010;365:1717–1733.

18. Zhang Z, Fye S, Borecki IB, Rader JS. Polymorphisms in immune mediators associate with risk of cervical cancer. Gynecol Oncol Oct 2014;135:69–73.

19. Dennis G, Jr., Sherman BT, Hosack DA, Yang J, Gao W, Lane HC, Lempicki RA. DAVID: Database for Annotation, Visualization, and Integrated Discovery. Genome Biol 2003;4:P3.

20. Tomomori S, Nakano Y, Ochi H, et al. Maintenance of low inflammation level by the ZFHX3 SNP rs2106261 minor allele contributes to reduced atrial fibrillation recurrence after pulmonary vein isolation. PLoS One 2018;13:e0203281.

21. Rabbani B, Khorgami M, Dalili M, Zamani N, Mahdieh N, Gollob MH. Novel cases of pediatric sudden cardiac death secondary to TRDN mutations presenting as long QT syndrome at rest and catecholaminergic polymorphic ventricular tachycardia during exercise: The TRDN arrhythmia syndrome. Am J Med Genet A Nov 2021;185:3433–3445.

22. Loggia ML, Bushnell MC, Tetreault M, Thiffault I, Bherer C, Mohammed NK, Kuchinad AA, Laferriere A, Dicaire MJ, Loisel L, Mogil JS, Brais B. Carriers of recessive WNK1/HSN2 mutations for hereditary sensory and autonomic neuropathy type 2 (HSAN2) are more sensitive to thermal stimuli. J Neurosci Feb 18 2009;29:2162–2166.

23. Gillespie D, Fill M. Pernicious attrition and inter-RyR2 CICR current control in cardiac muscle. J Mol Cell Cardiol May 2013;58:53–58.

24. Eisner DA, Caldwell JL, Kistamas K, Trafford AW. Calcium and Excitation-Contraction Coupling in the Heart. Circ Res Jul 7 2017;121:181–195.

25. Song L, Alcalai R, Arad M, Wolf CM, Toka O, Conner DA, Berul CI, Eldar M, Seidman CE, Seidman JG. Calsequestrin 2 (CASQ2) mutations increase expression of calreticulin and ryanodine receptors, causing catecholaminergic polymorphic ventricular tachycardia. J Clin Invest Jul 2007;117:1814–1823.

26. Nikolaienko R, Bovo E, Zima AV. Redox Dependent Modifications of Ryanodine Receptor: Basic Mechanisms and Implications in Heart Diseases. Front Physiol 2018;9:1775.

27. Chagula DB, Rechcinski T, Rudnicka K, Chmiela M. Ankyrins in human health and disease - an update of recent experimental findings. Arch Med Sci 2020;16:715–726.

28. Missiaen L, Dode L, Vanoevelen J, Raeymaekers L, Wuytack F. Calcium in the Golgi apparatus. Cell Calcium May 2007;41:405–416.

29. Knollmann BC, Chopra N, Hlaing T, et al. Casq2 deletion causes sarcoplasmic reticulum volume increase, premature Ca2+ release, and catecholaminergic polymorphic ventricular tachycardia. J Clin Invest Sep 2006;116:2510–2520.

30. Denegri M, Bongianino R, Lodola F, et al. Single delivery of an adeno-associated viral construct to transfer the CASQ2 gene to knock-in mice affected by catecholaminergic polymorphic ventricular tachycardia is able to cure the disease from birth to advanced age. Circulation Jun 24 2014;129:2673–2681.

31. Hwang HS, Hasdemir C, Laver D, Mehra D, Turhan K, Faggioni M, Yin H, Knollmann BC. Inhibition of cardiac Ca2+ release channels (RyR2) determines efficacy of class I antiarrhythmic drugs in catecholaminergic polymorphic ventricular tachycardia. Circ Arrhythm Electrophysiol Apr 2011;4:128–135.

32. Koenig SN, Mohler PJ. The evolving role of ankyrin-B in cardiovascular disease. Heart Rhythm Dec 2017;14:1884–1889.

33. Zhai C, Cong H, Liu Y, Zhang Y, Liu X, Zhang H, Ren Z. Rs7193343 polymorphism in zinc finger homeobox 3 (ZFHX3) gene and atrial fibrillation: an updated meta-analysis of 10 case-control comparisons. BMC Cardiovasc Disord Jun 26 2015;15:58.

34. Tsai CT, Hsieh CS, Chang SN, Chuang EY, Juang JM, Lin LY, Lai LP, Hwang JJ, Chiang FT, Lin JL. Next-generation sequencing of nine atrial fibrillation candidate genes identified novel de novo mutations in patients with extreme trait of atrial fibrillation. J Med Genet Jan 2015;52:28–36.

